# Pregnancy Loss and Cardiovascular Disease: A Nationwide Cohort Study

**DOI:** 10.1101/2021.01.04.21249212

**Authors:** Anders P. Mikkelsen, Pia Egerup, Astrid M. Kolte, David Westergaard, Christian Torp-Pedersen, Henriette S. Nielsen, øjvind Lidegaard

## Abstract

**Objectives:** To examine how pregnancy loss influences the risk of cardiovascular disease later in life.

**Design:** Prospective historical cohort study.

**Setting:** Danish nationwide health registries.

**Participants:** All Danish women with a recorded pregnancy from 1977 to 2017.

**Main outcome measures:** Venous thromboembolism, myocardial infarction, or ischemic stroke.

**Results:** In this two-part study, part one evaluated the 20-year absolute risk of cardiovascular disease from age 40 among 596,699 women with a full registered reproductive history. Adjusting for calendar year, diabetes, autoimmune disease, live births, and education, the absolute risk of an outcome after 0 and ≥4 pregnancy losses, respectably was: venous thromboembolism 3.0% (95% CI 2.8 to 3.2%) and 5.0% (3.4 to 6.8%); myocardial infarction 1.5% (1.4 to 1.6%) and 2.4% (1.4 to 3.6%); ischemic stroke 2.0% (1.9 to 2.1%) and 2.6% (1.5 to 3.6%). Prior stillbirth increased the absolute risk of later venous thromboembolism by 1.1% (0.2 to 2.3%); myocardial infarction by 1.1% (0.3 to 2.0%).

In study part two, we included 966,490 women from first pregnancy in a time-dependent Cox regression model. Adjusted for confounders, each additional pregnancy loss increased the hazard ratio of venous thromboembolism 1.10 (95% CI 1.07 to 1.13); myocardial infarction 1.12 (1.07 to 1.18); and ischemic stroke 1.10 (1.06 to 1.14). Stillbirth was strongly associated with myocardial infarction before age 40, adjusted hazard ratio of 4.60 (2.65 to 8.00).

**Conclusion:** Pregnancy loss was associated with later venous thromboembolism, myocardial infarction, and ischemic stroke. The absolute and relative risk of outcomes increased in a dose-response manner with increasing numbers of prior pregnancy losses. Stillbirth was strongly associated with myocardial infarction before age 40.

## Introduction

The leading cause of death worldwide is cardiovascular disease, accounting for around 31% of global deaths.[1] During the past decades the incidence of cardiovascular mortality and morbidity in the United States and Australia has decreased for almost all age groups, except among young women where the risk has either stagnated or increased.[2–4] Further, young women have longer hospital stays, and higher short-term mortality after myocardial infarctions than young men.[5,6] Existing risk stratification methods have proven less useful in young adults,[7] and clinicians often feel less prepared in assessing risk of cardiovascular disease in women.[8]

In order to identify women at risk of cardiovascular disease and further reduce the cardiovascular burden for young and middle-aged women, it is essential to evaluate the effect of new predictive factors. The reproductive profile is easily assessable and possibly an important indicator for at-risk women.[9] A first step in evaluating the applicability of pregnancy history as a marker for later cardiovascular disease, is large scale investigations into the strength of association. This could facilitate development of new risk stratification schemes and early prevention strategies, such as lifestyle modifications and prophylactic medication, for women not identified by traditional risk factors. Previous studies examining the effect of pregnancy loss on later cardiovascular disease, have shown ambiguous results, and have not presented clinically applicable absolute risk estimates. Further, they have not assessed the endpoint of venous thromboembolism, although this disease is common, underdiagnosed, and can lead to severe morbidity and death. [10–16] Although the mechanism behind the link between adverse pregnancy outcomes and later cardiovascular disease is unknown, systemic damage to the inner cell layer of vessel walls, termed endothelial dysfunction, is studied as a possible common pathway for both adverse pregnancy events and later cardiovascular disease.[17]

The aim of our study was to examine the effect of pregnancy loss, recurrent pregnancy loss, and stillbirth on later risk of venous thromboembolism, myocardial infarction, and ischemic stroke, to identify female risk factors for cardiovascular disease. For clinical interpretability of results, risk would be estimated in both absolute and relative measures using nationwide registers.

## Methods

### Data sources

Healthcare in Denmark is universal and free of charge. Data from a wide variety of healthcare services are collected in registries for administrative and research purposes. Using these nationwide registries, we extracted information on pregnancies, diseases and demographics for all Danish women between 1973 and 2017 with at least one registered pregnancy. The main data sources used were: (i) the Danish Central Persons Registry, containing date of birth and death, since 1968;[18] (ii) the Danish Medical Births Registry, containing data on live and stillbirths since 1973;[19] (iii) the Danish National Health Registry containing data on hospital discharge diagnosis codes since 1977;[20] (iv) the Danish National Prescription registry containing data on dispensed prescription medication since 1995,[21] (v) the Danish Cancer Registry, containing diagnoses of cancer, available since 1942.[22] A summary of registers used is listed in eTable 1. Search strings used to extract data from registries can be found in eTable 2. For diagnoses extracted from the National Health Registry, the admission date of primary and secondary diagnoses was used, and emergency room contacts were not used.

### Identification of unique pregnancies

Live births and stillbirths are registered in the Medical Births Registry. From 1973 to 1996 these events were filled manually for each birth. From 1997 data on births were extracted from the National Health Registry. In Denmark, the gestational threshold for a stillbirth to be reported in the Medical Births Registry, was 28 weeks until 2004, and 22 weeks hereafter.[19] Early pregnancy complications such as miscarriage, blighted ovum, ectopic pregnancy, and molar pregnancy were extracted from the National Health Registry. As these pregnancies can lead to multiple hospital visits during clinical care, a set of restriction periods were made to ensure that each pregnancy was only counted once: (i) at least 90 days between two early pregnancy complications, (ii) at least 154 days (22 weeks) between an early pregnancy complication and a succeeding delivery date of a live or stillbirth. (iii) At least 60 days between the delivery date of a live or stillborn and a succeeding early pregnancy complication. (iv) At least 154 days between a live or stillbirth and a succeeding live or stillbirth. In the rare event two pregnancies were overlapping, the first was kept.

### Study design

The study was conducted as a historical prospective cohort study.

Pregnancy loss was defined as a diagnosis code of miscarriage, spontaneous abortion, missed abortion, blighted ovum, or stillbirth. Three different pregnancy loss exposures were considered: (i) The number of pregnancy losses: 0, 1, 2, 3, or ≥4. (ii) Recurrent pregnancy loss, defined as either three consecutive registered pregnancy losses, or a diagnosis of recurrent pregnancy loss. *Primary recurrent pregnancy loss* was defined as women with no deliveries before a series of three or more consecutive pregnancy losses, and *secondary recurrent pregnancy loss* was defined as women with one or more births before the consecutive series of pregnancy losses. (iii) Stillbirth was also studied as an independent exposure.

Outcomes were defined as the first hospital contact, or the underlying cause of death of the following:(i) venous thromboembolism, including deep vein thrombosis, and pulmonary embolism (superficial thrombosis was not considered a valid outcome); (ii) myocardial infarction; (iii) ischemic stroke (transitory ischemic attack was not considered a valid outcome). Immigrants to Denmark were eligible for inclusion if they immigrated to Denmark before 20 years of age.

The study consisted of two separate designs in order to calculate both absolute and relative estimates: The first part evaluated absolute risk with a landmark approach at age 40 using Cause-specific Cox regression and G-formula;[16] the second part evaluated hazard ratios using time-dependent Cox regression.[17]

### Study population, study part one

In order to calculate 20-year absolute risks of outcomes, all women born from 1957 through 1977 with a registered pregnancy, were eligible for inclusion. Inclusion was based on a landmark at age 40.

Women were stratified in exposure groups according to pregnancy losses occurring before age 40. Women with a history of venous thromboembolism, myocardial infarction, ischemic stroke, thrombophilia, cancer, severe cardiovascular disease, emigration over six months, emigration with no return date, or death before inclusion were excluded.

### Statistical analyses, study part one

Risk time started at included women’ s 40^th^ birthday and ended at first occurrence of (i) an outcome of interest, (ii) death from other causes, resulting in a competing risk event. (iii) Emigration over six months, or end of follow-up (31^st^ of December 2017) resulting in censoring. Two Cox proportional hazards regression models were fit, one for the cause-specific outcome, and one for the competing outcome of death from other causes. Estimates were combined using G-formula to estimate the absolute risks.[23–25] The horizon for risk predictions was set to 20 years, corresponding to the probability for a 40 year old women to experience an event within the next 20 years, reported in percent. Absolute risks and risk differences were shown in a forest plot alongside 95% confidence intervals, calculated using 1,000 bootstrap samples.

### Study population, study part two

In this design women born from 1957 and onwards, with a registered pregnancy were eligible for inclusion. Women with a history of venous thromboembolism, myocardial infarction, ischemic stroke, thrombophilia, cancer, severe cardiovascular disease, or emigration over six months before first pregnancy were excluded. Women were included at first pregnancy after 1^st^ of January 1977 and followed until the earliest occurrence of the following: (i) venous thromboembolism, myocardial infarction, or ischemic stroke, resulting in an event for the respective outcome, or (ii) thrombophilia, malignant cancer, severe cardiovascular disease, emigration over six months, emigration with no return date, or end of follow up 31^st^ of December 2017, resulting in censoring.

### Statistical analyses, study part two

Included women were followed in a time-dependant manner, with maternal age as the underlying time axis. Each woman’ s risk time was split during follow-up, to account for covariates changing. The following occurrences caused the risk time to be split: (i) pregnancy loss, (ii) recurrent pregnancy loss, (iii) stillbirth, (iv) live birth, (v) diabetes, (vi) obtainment of a bachelor’ s degree, (vii) diagnosis of rheumatoid arthritis, (viii) diagnosis of systemic lupus erythematosus, and (ix) total risk time surpassed a 5 year interval. Women could increase exposure level to pregnancy loss during follow up, but not revert. Estimates were obtained using the Cox proportion hazards model, and the Cox proportional hazards assumption was tested by plotting the scaled Schoenfeld residuals against time, with acceptable horizontal linearity for all predicter variables, and no clear pattern with time. Estimates and 95% confidence intervals were displayed in forest plots.

### Sensitivity analysis, study part two

A sensitivity analysis was carried out for exposure to stillbirth and outcome of myocardial infarction. In this analysis women were censored as above or at age 40, whichever came first.

### Covariates

Covariates to adjust for in both study parts, were chosen after considering a causal graph (eFigure 1) of known and suspected confounders.[26] Analyses were adjusted for (i) calendar year using a spline with 3 degrees of freedom (ii) the number of live births using a spline with 3 degrees of freedom, (iii) diabetes, (iv) rheumatoid arthritis, (v) systemic lupus erythematosus, and (vi) educational level above or below bachelor degree, (vii) maternal age, as maternal age in years was used as the underlying time axis.

Hypertension was defined as at least two hospital admissions with a diagnosis of hypertension or fulfilling prescriptions of at least two classes of anti-hypertensive drugs (Classes: i. Adrenergic antagonist, ii. Diuretic, iii. Vasodilator, iv. Beta blocker, v. Calcium Antagonist, vi. Renin–Angiotensin– Aldosterone System Inhibitors). Diabetes included diagnoses for gestational diabetes and was defined as at least two hospital contacts with a diagnosis or fulfilling at least two prescription of an anti-diabetic drug. Thrombophilia, rheumatoid arthritis and systemic lupus erythematosus, and polycystic ovarian syndrome was defined as a relevant hospital diagnosis.

All data management and statistical analysis was performed using *R* Version 3.6.3.[27] Cox proportional hazards models were calculated using the *survival* package version 3.1-11.[28] Absolute risks were calculated using the *riskRegression* package version 2020.02.05.[29] Plots were produced using the *ggplot2* package version 3.3.2.[30] Causal graphs and minimal sufficient adjustment sets were obtained using the *dagitty* package version 0.3-0.[31]

### Patient and public involvement

Patients contributed data to the current study during hospital contacts and when purchasing prescription drugs, but were otherwise not involved in the research process, in forming the research question, in the design of the study, in the choice of outcome measures, and conduct of the study.

## Results

Study part one included 596,699 women, with 1,717,643 pregnancies before age 40. As seen in Figure 1, exposure to 3 and ≥4 registered pregnancy losses before age 40 occurred for 4,715 (0.8%) and 1,664 (0.3%) women, respectively. Exposure to consecutive pregnancy losses, termed primary recurrent pregnancy losses and secondary recurrent pregnancy loss, occurred for 2,940 (0.5%) and 3,281 (0.5%) women, respectively. Baseline characteristics at age 40, seen in Table 1, illustrate that women with increasing numbers of pregnancy losses, more often had no live births and more seldom obtained a bachelor’ s degree. Among women with ≥4 pregnancy losses, 9.5% had no live births at age 40, compared to 3.8% among women with no pregnancy losses. The proportion of women with comorbidities such as diabetes, polycystic ovarian syndrome, prior preeclampsia, or lupus erythematosus, increased with the number of prior pregnancy losses. The share of women who were smokers and/or had a high body mass index also increased with increasing number of pregnancy losses. Information about smoking was available for 75.0% and body mass index was available for 24.8% of women in study part one.

**Figure 1.**
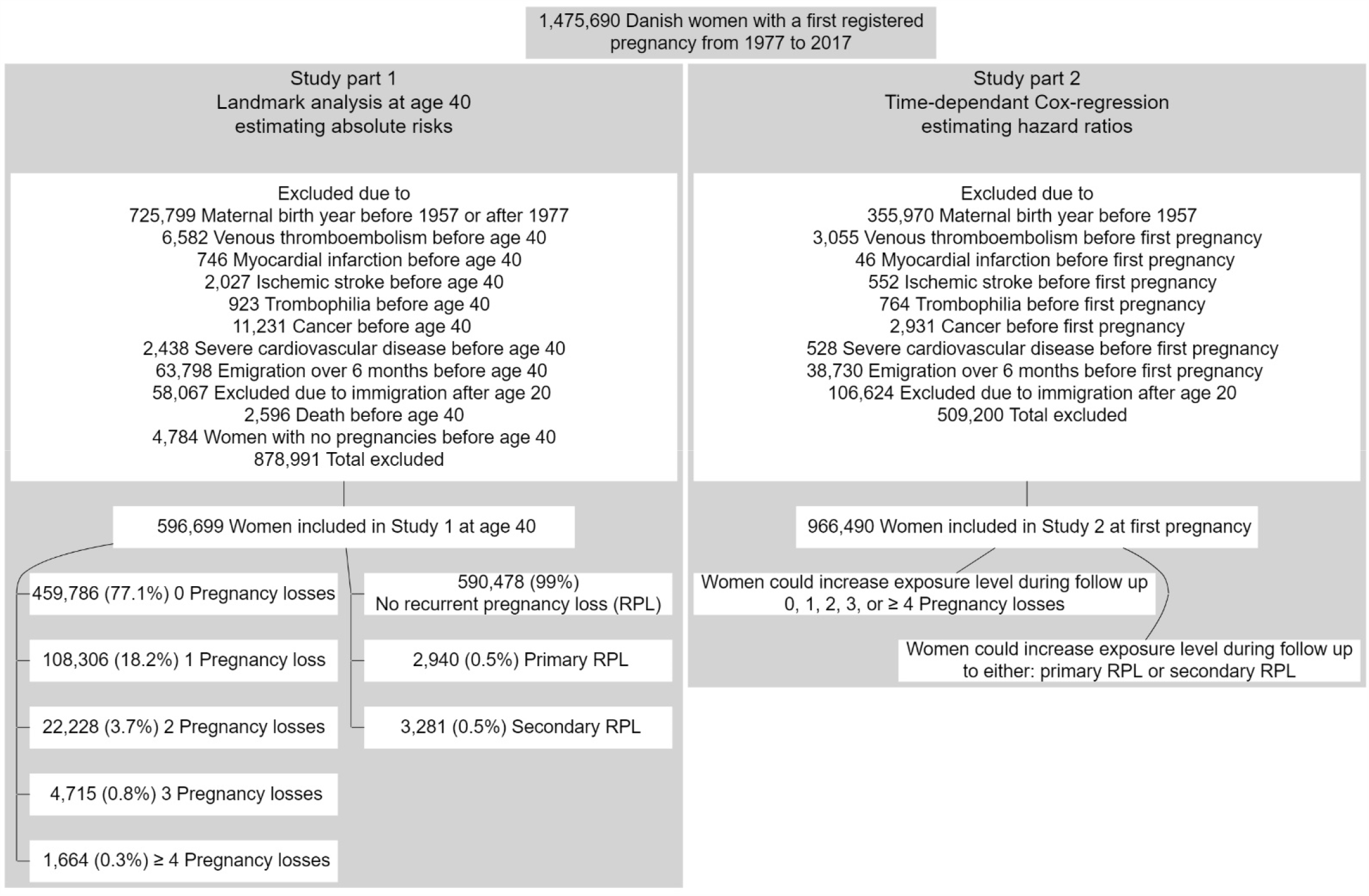
Inclusion and exclusion criteria.

**Table 1.** Baseline characteristics for woman at age 40, included in study part 1. Women were stratified by number of pregnancy losses. *Diabetes included diabetes type 1, type 2, and gestational diabetes. **Information about smoking habits was available for 75.0%, and body mass index was available for 24.8% of the cohort.

| Ever pregnant women, stratified by number of pregnancy losses before age 40 |  |  |  |  |  |
| --- | --- | --- | --- | --- | --- |
|  | 0 | 1 | 2 | 3 | ≥ 4 |
| n | 459,786 | 108,306 | 22,228 | 4,715 | 1,664 |
| Number of live births, n (%) |  |  |  |  |  |
| 0 | 17,417 ( 3.8) | 5,639 ( 5.2) | 1,096 ( 4.9) | 235 ( 5.0) | 158 ( 9.5) |
| 1 | 8,9421 (19.4) | 16,099 (14.9) | 3,395 (15.3) | 781 (16.6) | 339 (20.4) |
| ≥ 2 | 35,2948 (76.8) | 86,568 (79.9) | 17,737 (79.8) | 3,699 (78.5) | 1,167 (70.1) |
| Stillbirth, n (%) | 0 ( 0.0) | 1,566 ( 1.4) | 601 ( 2.7) | 163 ( 3.5) | 65 ( 3.9) |
| Gravidity (mean (SD)) | 2.54 (1.20) | 3.75 (1.42) | 4.93 (1.58) | 6.07 (1.70) | 7.43 (2.13) |
| Bachelor degree or higher, n (%) | 133,553 (29.0) | 32,172 (29.7) | 6,518 (29.3) | 1,368 (29.0) | 394 (23.7) |
| Diabetes*, n (%) | 1,1649 ( 2.5) | 3,418 ( 3.2) | 881 ( 4.0) | 220 ( 4.7) | 94 ( 5.6) |
| Polycystic ovarian syndrome, n (%) | 4,884 ( 1.1) | 1,454 ( 1.3) | 327 ( 1.5) | 85 ( 1.8) | 30 ( 1.8) |
| Hypertension, n (%) | 17,714 ( 3.9) | 4,325 ( 4.0) | 971 ( 4.4) | 180 ( 3.8) | 74 ( 4.4) |
| Rheumatoid arthritis, n (%) | 2,650 ( 0.6) | 627 ( 0.6) | 109 ( 0.5) | 35 ( 0.7) | 11 ( 0.7) |
| Systemic lupus erythematosus, n (%) | 507 ( 0.1) | 108 ( 0.1) | 32 ( 0.1) | 12 ( 0.3) | 7 ( 0.4) |
| Preeclampsia/ eclampsia n (%) | 26,042 ( 5.7) | 6,428 ( 5.9) | 1,400 ( 6.3) | 345 ( 7.3) | 121 ( 7.3) |
| Smoking, n (%)** | 79,766 (23.5) | 20,987 (24.8) | 4,725 (26.4) | 1,072 (28.0) | 382 (30.1) |
| Body mass index n (%)** |  |  |  |  |  |
| <18.5 | 3,408 ( 3.1) | 888 ( 3.1) | 205 ( 3.2) | 56 ( 4.0) | 18 ( 4.2) |
| 18.5-29.9 | 93,119 (84.1) | 24,347 (83.7) | 5,298 (82.5) | 1,111 (79.4) | 340 (78.7) |
| ≥ 30 | 14,136 (12.8) | 3,840 (13.2) | 921 (14.3) | 232 (16.6) | 74 (17.1) |

Estimated 20-year absolute risks of all outcomes at age 40, increased with each additional pregnancy loss as seen in Figure 2. Venous thromboembolism was the most frequent outcome with 6,978 events followed by 5,488 ischemic strokes, and 3,654 myocardial infarctions. The median follow-up time for all outcomes was 10.8 years (interquartile range 5.5 to 15.6 years). The 20-year adjusted absolute risk of venous thromboembolism was significantly increased after ≥4 pregnancy losses, secondary recurrent pregnancy loss, and after stillbirth. The absolute risk of venous thromboembolism was 3.0% (95% confidence interval 2.8 to 3.2%) for women with no pregnancy losses and increased to 5.0% (3.4 to 6.8%) after ≥4 prior pregnancy losses, risk difference 2.0% (0.5 to 3.8%). The absolute risk of myocardial infarction was significantly increased after two pregnancy losses, secondary recurrent pregnancy loss, or stillbirth, e.g. the risk was 1.5% (1.4 to 1.6%) after no stillbirths, and rose to 2.6% (1.8 to 3-5%) after a stillbirth, increasing 1.1% (0.3 to 2.0%). For the outcome of ischemic stroke, primary and secondary recurrent pregnancy loss, and prior stillbirth increased the absolute risk significantly, e.g. after no recurrent pregnancy loss the absolute risk of ischemic stroke was 2.0% (1.9 to 2.1%), increasing to 3.2% (2.3 to 4.2%) after primary recurrent pregnancy loss, a risk difference of 1.2% (0.3 to 2.2%).

**Figure 2.**
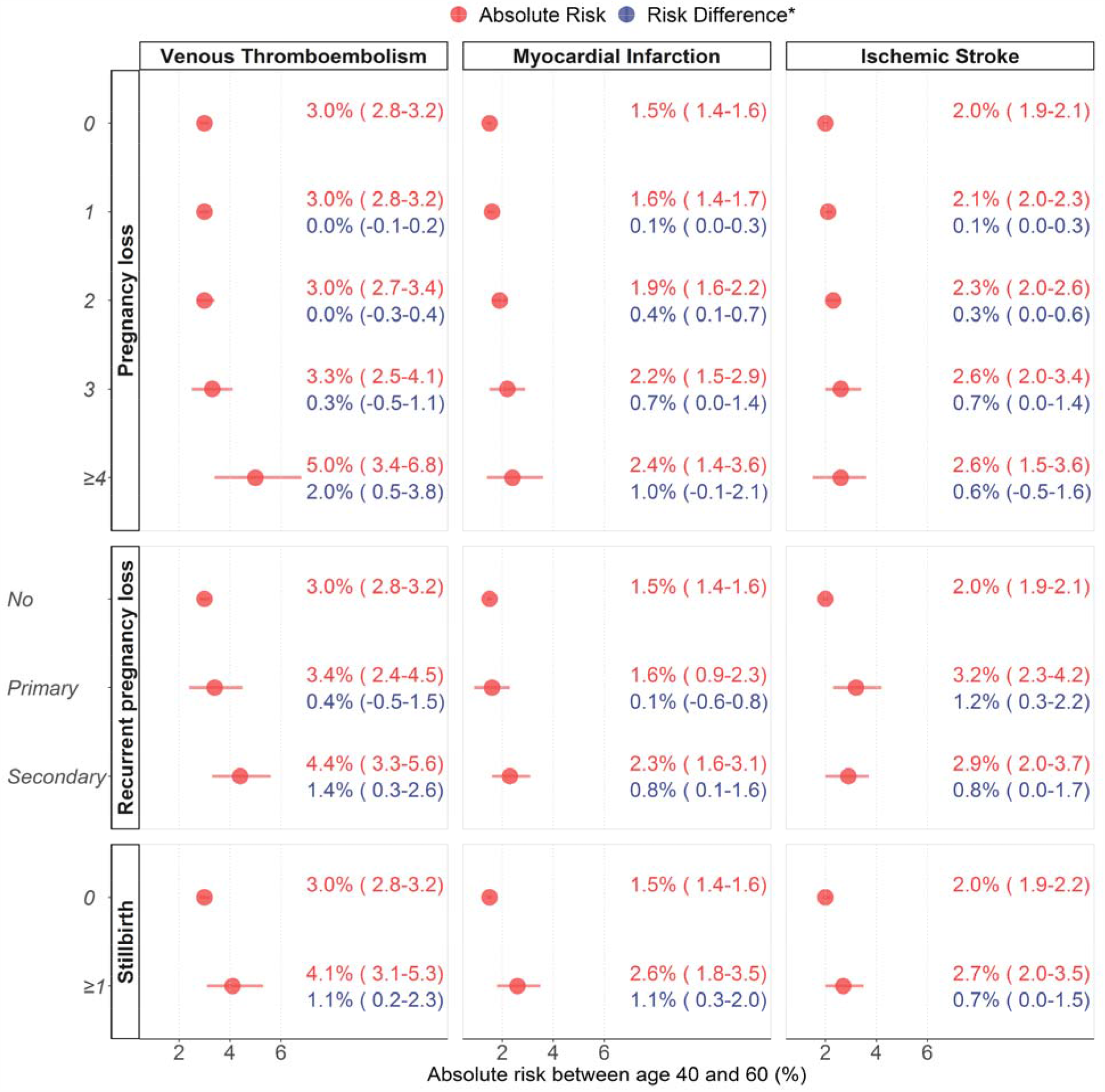
Results from study part one, estimating 20-year absolute risks of cardiovascular outcomes at age 40, stratified by number and type of pregnancy loss. Upper part: Exposure to increasing number of pregnancy losses. Middle part: exposure to either primary recurrent pregnancy loss, defined as three consecutive pregnancy losses, not preceded by a birth; or secondary recurrent pregnancy loss, preceded by a birth. Lower part: Exposure to one or more stillbirths. Models adjusted for calendar year, diabetes, rheumatoid arthritis, systemic lupus erythematosus, number of live births, and obtainment of bachelor’ s degree or higher. *Risk difference indicates difference in risk compared to reference. (Parentheses) show 95% confidence intervals.

The time-dependent model in study part two followed 966,490 women with a total of 2,575,503 pregnancies. In this study, the number of events experienced during follow-up was 12,077 venous thromboembolisms, 3,750 myocardial infarctions, and 6,745 ischemic strokes. The median follow-up time was 17.7 years (inter quantile range 8.4 to 27.0 years). The results were similar to study part one, as seen in Figure 3. Each additional pregnancy loss increased the hazard ratio of all three outcomes, compared to women with no pregnancy losses. The largest increase in hazard ratio for each outcome was; for myocardial infarction after stillbirth versus no stillbirth: adjusted hazard ratio 2.40 (1.83 to 3.16); for venous thromboembolism after ≥4 pregnancy losses versus 0 pregnancy losses: adjusted hazard ratio 1.75 (1.33 to 2.32); and for ischemic stroke after primary recurrent pregnancy loss versus no recurrent pregnancy loss: adjusted hazard ratio 1.62 (1.24 to 2.11). Each additional pregnancy loss (marginal effect) increased the hazard ratio of venous thromboembolism by 1.10 (1.07 to 1.13), myocardial infarction by 1.12 (1.07 to 1.18), and ischemic stroke by 1.10 (1.06 to 1.14) as seen in eTable 3.

**Figure 3.**
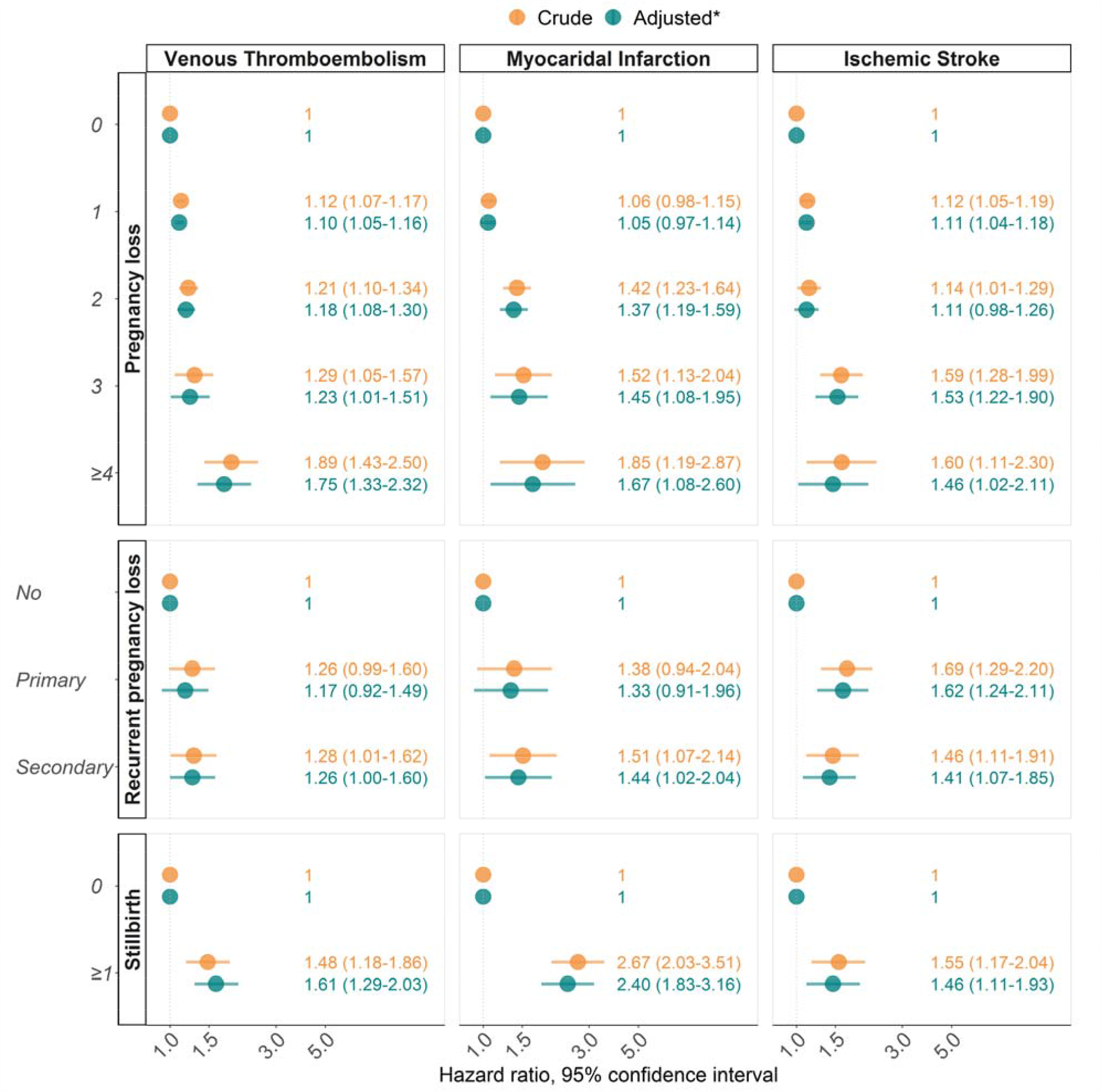
Results from time-dependent Cox regressions of study part two. Upper part: exposure to increasing numbers of pregnancy loss. Middle part: exposure to either primary recurrent pregnancy loss, defined as three consecutive pregnancy losses, not preceded by a birth; or secondary recurrent pregnancy loss, preceded by a birth. Lower part: exposure to stillbirth. Crude hazard ratios shown in orange. *Estimates in blue adjusted for: calendar year, maternal age, diabetes, rheumatoid arthritis, systemic lupus erythematosus, number of lives births, and obtainment of bachelor’ s degree or higher. (Parentheses) show 95% confidence intervals.

Stratifying the analysis for myocardial infarction after stillbirth by age, revealed women under age 40 to have an adjusted hazard ratio of 4.60 (2.65 to 8.00), compared to women without stillbirth (eTable 4).

## Discussion

In this nationwide cohort study including almost one million women, and 2.6 million pregnancies during a 41-year study period, pregnancy loss and stillbirth was associated with an increased absolute and relative risk of venous thromboembolism, myocardial infarction, and ischemic stroke. The risk increased after each pregnancy loss, in a dose-response relationship.

The study was methodically divided into two parts, to allow for evaluation of both absolute and relative effects of pregnancy loss on later cardiovascular disease. There were major similarities in results between the two study parts, however, also some differences. As the time-dependant model in study part two included women from their first pregnancy instead of at age 40, it included a 38.3% larger cohort and 28.6% more events, increasing precision in risk estimates and reducing confidence bands. The effect of even one or two pregnancy losses on outcomes was significant in part two, likely due to the increased power. By design only part one could evaluate clinically relevant risk absolute risk estimates and take the competing risk of death from other causes into account, which becomes highly relevant when following a cohort for up to 20 years. A risk estimate which was markedly different between the two study parts was the analysis of myocardial infarction after stillbirth. We therefor stratified this analysis on age, which revealed the hazard of myocardial infarction was especially pronounced before age 40.

### Strengths and weaknesses of the study

The major strength of this study is the size of the study population, length of follow-up, and the clinically confirmed endpoints.

The study was limited by data registered. Variables such as smoking status and body mass index commenced during follow-up and was only available for women who ever experienced a live or stillbirth. As body mass index and smoking were not part of the suggested minimal sufficient adjustment set of the causal graph (eFigure 1), this was not seen as major limitation.

A diagnosis of pregnancy loss was defined by diagnoses in the National Health Registry. These diagnoses have been shown to have a positive predictive value of 97% (92.7-99.5).[32] The health registry only contains hospital-treated pregnancy losses, and we have therefore likely underestimated the true number, as those managed at home, by general practitioners, and gynaecologists in private practice, are not registered. Thrombophilia was likely underdiagnosed, but not under-reported, as during much of follow-up, thrombophilia evaluation was not carried out until after a thromboembolic event or evaluation for recurrent pregnancy loss. A hospital diagnosis of venous thromboembolism has been shown to have a positive predictive value of 91.2%,[33] while validation of a hospital diagnosis of myocardial infarction has shown a positive predictive value of 93.6% and sensitivity of 77.6%.[34] For stroke the corresponding positive predictive value was 97.0%.[35] No studies have to our knowledge validated the equivalent diagnoses in the cause of death register, however very few of outcome diagnoses were only identified in the cause of death register (eTable 5).

### Strengths and weaknesses in relation to other studies, discussing important differences in results

Results from studies, examining the association between pregnancy loss, and later myocardial infarction and ischemic stroke have shown ambiguous results,[10,11,14] and meta-analyses have as a result concluded that there is not enough evidence to draw meaningful conclusions about the possible link, although prior stillbirth is increasingly accepted to be a risk factor for later cardiovascular disease.[9,36]

To our knowledge, no studies have evaluated the outcome of venous thromboembolism, and further no studies have presented clinically meaningful absolute risk estimates. Adjustment of confounders varied among studies examining the risk of myocardial infarction and stroke after pregnancy loss. In the largest cohort study to date, risk estimates were adjusted for current age, calendar period, and number of live births.[14]. Our study supports the main conclusions from the study, although there are some discrepancies in our findings, e.g. the study found a decreased incidence rate ratio for myocardial infarction after three miscarriages compared to none.

The rare autoimmune disease antiphospholipid syndrome is linked to both pregnancy loss and thrombotic events, and is present in 5-15% of patients with recurrent pregnancy loss.[37,38] Possibly other immunological disturbances influence both pregnancy loss and later risk of thrombosis. Our findings suggest that women with secondary recurrent pregnancy loss are at an increased risk of venous thrombosis, and myocardial infarction, while women with primary recurrent pregnancy loss have an increased risk of later ischemic stroke, however, the increase in risk was not substantially different than for women with multiple non-recurrent losses or stillbirth.

### Meaning of the study: possible explanations and implications for clinicians and policymakers

A woman’ s pregnancy profile could provide clinicians with a valuable and easily assessable tool containing information about future risk of venous thromboembolism, myocardial infarction, and ischemic stroke. Our study provides easily interpretable 20-year absolute risks of outcomes from age 40, based on history of pregnancy loss. For implementation in current risk stratification schemes, further investigations into the added effect of including pregnancy loss history, and the effect of lifestyle modification and medical intervention strategies.

### Unanswered questions and future research

The mechanisms behind the associations cannot be directly deduced from our results. The pregnancy losses could themselves have increased the risk of outcomes, possibly through specific physiological changes caused by the pregnancy loss. Alternatively, a genetic, metabolic, vascular, or immune precursor exists, which is associated with both pregnancy loss and cardiovascular disease, possibly through endothelial dysfunction. Further, the large increase in risk of myocardial infarction among women under 40 warrants further investigation.

## Conclusion

Pregnancy loss and stillbirth was associated with later venous thromboembolism, myocardial infarction, and ischemic stroke. Each additional pregnancy loss increased the absolute and relative risk of venous and arterial thrombosis in a dose-response relationship. The hazard ratio of myocardial infarction after a stillbirth was especially elevated before age 40.

## Data Availability

Data is not publicaly available. Only aggregated data can be exported for research purposes after relevant approval.

## Contributors and sources

APM, PE, AMK, HSN, OL conceptualized the study. OL obtained access to data. APM performed data management, statistical analyses, and wrote the initial version of the manuscript. CTP provided important insight during data analysis. All authors participated in interpretation of data and revised the manuscript critically. All authors had full access to all the data in the study and can take full responsibility for the integrity and the accuracy of the data analysis. APM is the guarantor. The corresponding author APM attests that all authors meet authorship criteria and that no others meeting the criteria have been omitted.

## Approval

Data was provided from the Danish Health Data Authority under license FSEID-00003879. Permission was obtained from the Regional Data Protection Agency (P-2020-217). No ethical approval is needed for registry-based research.

## Transparency statement

The lead author affirms that the manuscript is an honest, accurate, and transparent account of the study being reported; no important aspects of the study have been omitted; and discrepancies from the originally planned study are explained.

## Data sharing

No additional data are available.

## Supplementary Material

**eTable 1.** Nationwide registries used in study.

| Register | Available since | Data used |
| --- | --- | --- |
| Cancer Registry | 1942 | Date of cancer diagnosis |
| Central Persons Registry | 1968 | Date of birth and death |
| Cause of Death Registry | 1969 | Primary cause of death |
| Induced Abortion Registry | 1970 | Date of induced abortion |
| Immigration and emigration Registry | 1973 | Immigration and emigration dates |
| Medical Births Registry | 1973 | Date of delivery, and pregnancy outcome |
| National Patient Registry | 1977 | Hospital discharge diagnosis and date |
| Education Registry | 1981 | Date of educational degree |
| National Prescription Registry | 1995 | Medication dispensed at Danish pharmacies |

**eTable 2.** Search strings used for diagnosis definitions. Only primary and secondary diagnosis codes were used. Emergency room contacts were not used. ICD-8: International Classification of Diseases 8th revision. ICD-10: 10th revision. ATC: Anatomical Therapeutic Chemical Classification. [Brackets]: Indicate an interval of codes, both characters included. All subdiagnoses were included, e.g. I26 Pulmonary embolism included I26.0 Pulmonary embolism with acute cor pulmonale.

| Definition | ICD-8 | ICD-10 | Other |
| --- | --- | --- | --- |
| Live birth |  |  | Live birth in Medical Births Register |
| Stillbirth |  |  | Stillbirth in Medical Births Register |
| Early pregnancy loss | 6346, 643[8-9], 6451 | O020, O021, DO03 |  |
| Extrauterine pregnancy | 631[0-6], 63199 | O00 |  |
| Molar pregnancy | 63190, 63429, 6450, 63460 | D392, O01 |  |
| Recurrent Pregnancy Loss (RPL) | Y6439,6430 | N96, O262 | or 3 consecutive diagnoses of early pregnancy loss. Primary RPL not preceded by a live birth |
| Venous Thromboembolism | 450, 45100, 45108, 45199, 452, 45302, 67101, 6710[8-9], 673 | I26, I80[1-3], I80[8-9], I81, I82[2-9], O082, O087, O223, O225, O871, O873, O87[8-9], O88[0-2]) |  |
| Myocardial Infarction | 410 | I2[1-2] |  |
| Stroke | 43[3-4], 436 | I6[3-4] |  |
| Thrombophilia |  | D68[5-6] |  |
| Other CVD | 41[1-4], 431 | I2[3-5], I6[1-2] |  |
| Diabetes/gestational diabetes | 249, 250, 63474 | E1[0-1], E14, O24, | 2 diagnosis codes and/or 2 fulfillments of drug class ATC: A10 |
| Polycystic ovarian syndrome | 25690 | E282, L680 |  |
| Hypertension | 40[0-4], 4110, 4120, 4130, 4140, 63700 | I1[0-5] | 2 diagnosis codes and/or fulfillment of at least 2 types of bloodpressure lowering drugs:\n(i) Adrenergic antagonist ATC: C02[A-C], (ii) Diuretic C02DA, C03[A-B], C03[D-E], C03X, C07[B-D], C08G, C09BA, C09DA, C09XA52, C02L, (iii) Vasodilator C02DB, C02DD, C02DG, (iv) Beta Blocker C07[A-D], C07F, (v) Calcium Antagonist C07FB, C08, C09BB, C09DB, (vi) Renin-Angiotensin Inhibitor C09AA, C09BA, C09BB, C09CA, C09DA, C09DB, C09XA02, C09XA52 |
| Cancer |  |  | Diagnosis of malignant cancer in the Cancer register (not including non-melanoma skin cancer) |
| Preeclampsia/eclampsia | 6370[3-9], 63719, 76219, 76229 | O11, O14, O15 |  |
| Rheumatoid arthritis | 712[0-3], 69549 | M05, M06, M08 |  |
| Systemic lupus erythematosus | 73419 | M32 |  |

**eTable 3.** Events and risk time in study part 2. *Analysis adjusted for maternal age, calendar year, diabetes, systemic lupus erythematosus, rheumatoid arthritis, and educational level above or below bachelor’ s degree

| Outcome | Exposure | Exposure levels | Events | Risk time (years) | Crude | Adjusted* |
| --- | --- | --- | --- | --- | --- | --- |
| Venous Thromboembolism | Pregnancy loss | 0 | 9,334 | 14,038,290 | 1 | 1 |
| Venous Thromboembolism | Pregnancy loss | 1 | 2,152 | 2,729,391 | 1.12 (1.07-1.17) | 1.10 (1.05-1.16) |
| Venous Thromboembolism | Pregnancy loss | 2 | 444 | 495,439 | 1.21 (1.10-1.34) | 1.18 (1.08-1.30) |
| Venous Thromboembolism | Pregnancy loss | 3 | 97 | 99,354 | 1.29 (1.05-1.57) | 1.23 (1.01-1.51) |
| Venous Thromboembolism | Pregnancy loss | >=4 | 50 | 34,095 | 1.89 (1.43-2.50) | 1.75 (1.33-2.32) |
| Venous Thromboembolism | Pregnancy loss | marginal_effect | - | - | 1.12 (1.08-1.15) | 1.10 (1.07-1.13) |
| Venous Thromboembolism | Recurrent pregnancy loss | No | 11,941 | 17,256,157 | 1 | 1 |
| Venous Thromboembolism | Recurrent pregnancy loss | Primary | 67 | 71,751 | 1.26 (0.99-1.60) | 1.17 (0.92-1.49) |
| Venous Thromboembolism | Recurrent pregnancy loss | Secondary | 69 | 68,661 | 1.28 (1.01-1.62) | 1.26 (1.00-1.60) |
| Venous Thromboembolism | Stillbirth | 0 | 12,002 | 17,329,967 | 1 | 1 |
| Venous Thromboembolism | Stillbirth | >=1 | 75 | 66,602 | 1.48 (1.18-1.86) | 1.61 (1.29-2.03) |
| Myocardial Infarction | Pregnancy loss | 0 | 2,802 | 14,038,290 | 1 | 1 |
| Myocardial Infarction | Pregnancy loss | 1 | 690 | 2,729,391 | 1.06 (0.98-1.15) | 1.05 (0.97-1.14) |
| Myocardial Infarction | Pregnancy loss | 2 | 193 | 495,439 | 1.42 (1.23-1.64) | 1.37 (1.19-1.59) |
| Myocardial Infarction | Pregnancy loss | 3 | 45 | 99,354 | 1.52 (1.13-2.04) | 1.45 (1.08-1.95) |
| Myocardial Infarction | Pregnancy loss | >=4 | 20 | 34,095 | 1.85 (1.19-2.87) | 1.67 (1.08-2.60) |
| Myocardial Infarction | Pregnancy loss | marginal_effect | - | - | 1.14 (1.09-1.20) | 1.12 (1.07-1.18) |
| Myocardial Infarction | Recurrent pregnancy loss | No | 3,692 | 17,256,157 | 1 | 1 |
| Myocardial Infarction | Recurrent pregnancy loss | Primary | 26 | 71,751 | 1.38 (0.94-2.04) | 1.33 (0.91-1.96) |
| Myocardial Infarction | Recurrent pregnancy loss | Secondary | 32 | 68,661 | 1.51 (1.07-2.14) | 1.44 (1.02-2.04) |
| Myocardial Infarction | Stillbirth | 0 | 3,698 | 17,329,967 | 1 | 1 |
| Myocardial Infarction | Stillbirth | >=1 | 52 | 66,602 | 2.67 (2.03-3.51) | 2.40 (1.83-3.16) |
| Ischemic Stroke | Pregnancy loss | 0 | 5,092 | 14,038,290 | 1 | 1 |
| Ischemic Stroke | Pregnancy loss | 1 | 1,277 | 2,729,391 | 1.12 (1.05-1.19) | 1.11 (1.04-1.18) |
| Ischemic Stroke | Pregnancy loss | 2 | 267 | 495,439 | 1.14 (1.01-1.29) | 1.11 (0.98-1.26) |
| Ischemic Stroke | Pregnancy loss | 3 | 80 | 99,354 | 1.59 (1.28-1.99) | 1.53 (1.22-1.90) |
| Ischemic Stroke | Pregnancy loss | >=4 | 29 | 34,095 | 1.60 (1.11-2.30) | 1.46 (1.02-2.11) |
| Ischemic Stroke | Pregnancy loss | marginal_effect | - | - | 1.11 (1.07-1.15) | 1.10 (1.06-1.14) |
| Ischemic Stroke | Recurrent pregnancy loss | No | 6,638 | 17,256,157 | 1 | 1 |
| Ischemic Stroke | Recurrent pregnancy loss | Primary | 55 | 71,751 | 1.69 (1.29-2.20) | 1.62 (1.24-2.11) |
| Ischemic Stroke | Recurrent pregnancy loss | Secondary | 52 | 68,661 | 1.46 (1.11-1.91) | 1.41 (1.07-1.85) |
| Ischemic Stroke | Stillbirth | 0 | 6,694 | 17,329,967 | 1 | 1 |
| Ischemic Stroke | Stillbirth | >=1 | 51 | 66,602 | 1.55 (1.17-2.04) | 1.46 (1.11-1.93) |

**eTable 4.** Hazard ratios for the analysis on stillbirth and later risk of myocardial infarction before age 40 in study part two. Risk time in this analysis ended at age 40. *Analysis adjusted for maternal age, calendar year, diabetes, systemic lupus erythematosus, rheumatoid arthritis, and educational level above or below bachelor’ s degree.

| Outcome | Exposure levels | Events | Risk time (years) | Crude hazard ratio | Adjusted hazard ratio* |
| --- | --- | --- | --- | --- | --- |
| Myocardial infarction before age 40 | No stillbirth | 685 | 1,123,382 | 1 | 1 |
|  | Stillbirth | 13 | 34,639 | 5.4 (3.1-9.4) | 4.6 (2.7-8.0) |

**eTable 5.**
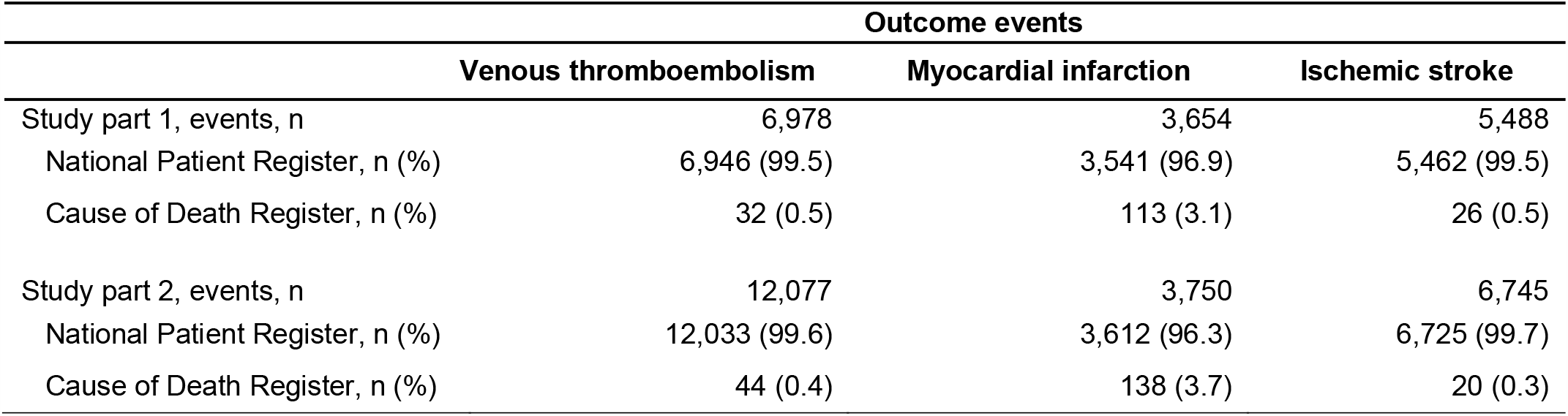
The number of events and the register source of these events.

|  | Outcome events |  |  |
| --- | --- | --- | --- |
|  | Venous thromboembolism | Myocardial infarction | Ischemic stroke |
| Study part 1, events, n | 6,978 | 3,654 | 5,488 |
| National Patient Register, n (%) | 6,946 (99.5) | 3,541 (96.9) | 5,462 (99.5) |
| Cause of Death Register, n (%) | 32 (0.5) | 113 (3.1) | 26 (0.5) |
| Study part 2, events, n | 12,077 | 3,750 | 6,745 |
| National Patient Register, n (%) | 12,033 (99.6) | 3,612 (96.3) | 6,725 (99.7) |
| Cause of Death Register, n (%) | 44 (0.4) | 138 (3.7) | 20 (0.3) |

**eFigure 1.**
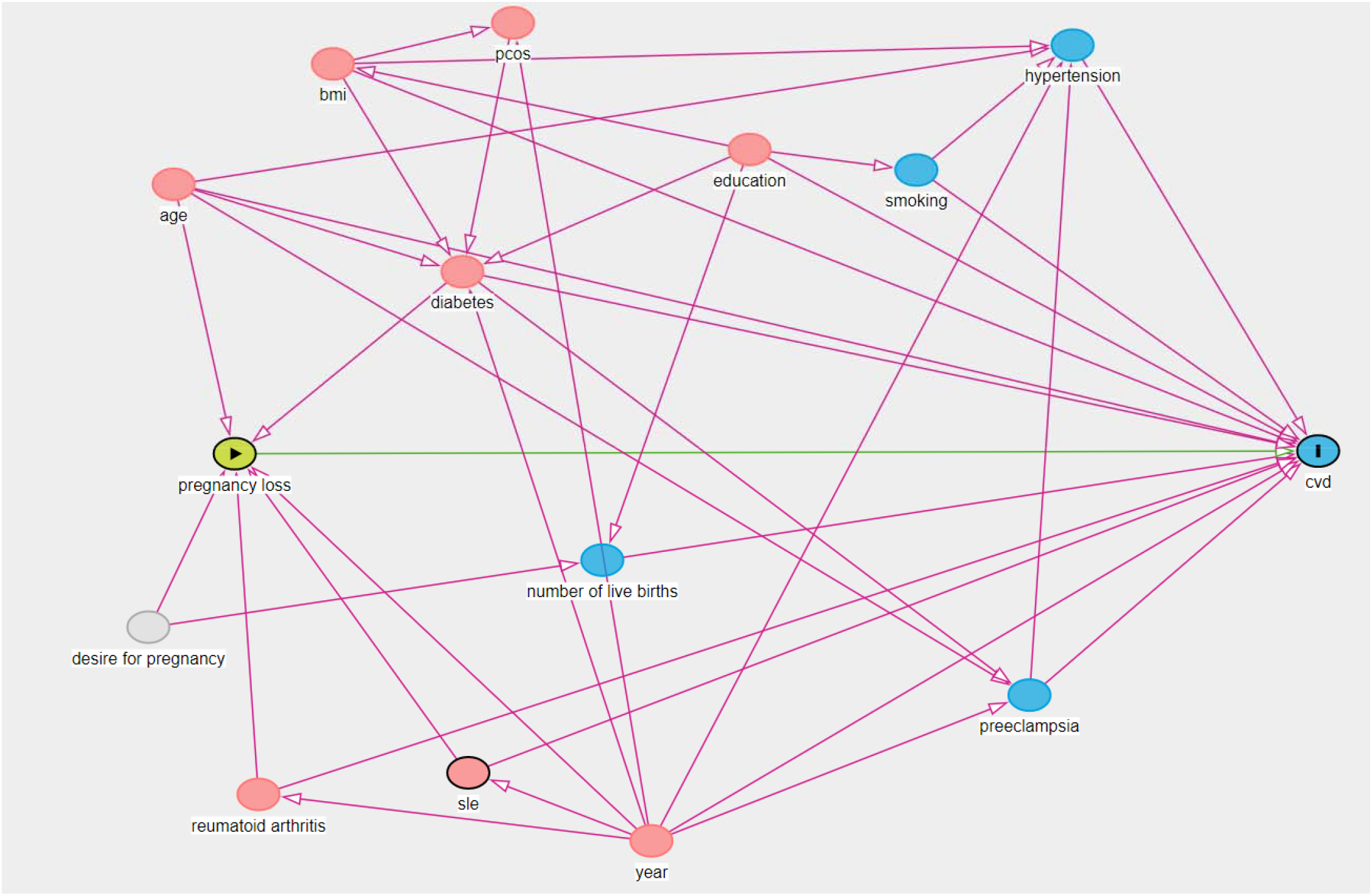
Directed Acyclic Graph. Green indicate exposure variable. “I” Indicates outcome variable. Red indicates measured covariates. Grey indicates unmeasured variables. CVD: cardiovascular disease. BMI: Body mass index. PCOS: polycystic ovarian syndrome. Minimal sufficient adjustment set for the direct and total effect: age, diabetes, education, number of live births, rheumatoid arthritis, year. Results obtained using dagitty^1^. ^1^Textor J, Zander B van der, Gilthorpe MS, et al. Robust causal inference using directed acyclic graphs: the R package ‘dagitty’. Int J Epidemiol 2016;45:1887–1894. doi:10.1093/ije/dyw341

